# The association of osteoarthritis with the risk of de novo inflammatory arthritis in patients receiving immune checkpoint inhibitors: a retrospective study

**DOI:** 10.64898/2026.01.28.26344880

**Authors:** Shiju Chen, Xingxing Zhu, Zhiqiang Zhang, Uma Thanarajasingam, Cynthia S. Crowson, Hu Zeng

## Abstract

**Objective:** Identifying risk factors enables stratification of patients’ susceptibility to inflammatory arthritis immune-related adverse events (IA-irAE). This retrospective study examines whether preexisting osteoarthritis (OA) increases the likelihood of *de novo* IA in patients treated with immune checkpoint inhibitors (ICIs).

**Methods:** The prevalence of OA among ICI–treated patients who developed IA-irAE, those who developed other types of irAEs but not IA (non-IA irAE), and those who did not develop any irAEs (non-irAE) were compared. Electronic medical records were reviewed to extract demographic, clinical and laboratory data. Group comparisons and logistic regression analyses were performed.

**Results:** 181 *de novo* IA-irAE patients, 140 non-IA irAE patients and 170 non-irAE patients were included. The prevalence of OA was significantly higher in the IA-irAE group (69%) than the non-IA irAE group (48%) and the non-irAE group (48%) (both p < 0.001). The IA-irAE group demonstrated a higher frequency of multisite OA, with predominant hand involvement (62%) than the non-IA irAE with OA group (13%) and the non-irAE with OA group (13%) (both p < 0.001). A family history of autoimmune disease (AID) (OR 2.03, 95% CI 1.02–4.05), preexisting OA (OR 2.88, 95% CI 1.85–4.52) and melanoma (OR 2.63, 95% CI 1.56–4.47) were identified as risk factors for the development of IA-irAE.

**Conclusions:** OA was more prevalent among ICI-treated patients developing IA-irAE than those who did not. Hand OA was the most common OA pattern in IA-irAE patients. Preexisting OA, melanoma and a family history of AID were risk factors for IA-irAE.

## INTRODUCTION

Immune checkpoint inhibitors (ICIs) have revolutionized cancer therapy; however, ICI therapy can over-activate the immune system, which leads to a spectrum of immune-related adverse events (irAEs) [1]. It has been reported that approximately 72% of the patients receiving PD-1 blockade, 59% in PD-L1 blockade therapies and up to 83% in those treated with anti–CTLA-4 blockade developed irAE with any grade [2], and the mean event rate of general irAEs of any grade was reported to be 40% in a recent systemic review [3]. IrAEs can involve nearly any organ system during or after ICI treatment and have been associated with improved oncologic outcomes [4, 5]. Since the first description of the clinical features of inflammatory arthritis (IA) irAE (IA-irAE), the spectrum of rheumatic irAEs has become increasingly recognized [6, 7]. De novo IA-irAE is the most common type of rheumatic irAEs with the prevalence of 3.8% [4]. The clinical manifestations of rheumatic irAEs warrant accurate diagnosis, comprehensive characterization and management by rheumatologists. Identifying the clinical risk factors for IA-irAE is therefore essential to better define and identify susceptible patient populations. To date, a limited number of clinical characteristics including the tumor type, autoimmune disease (AID) history have been proposed as risk factors for IA-irAE [8, 9].

A recent study indicated that preexisting osteoarthritis (OA) may be associated with an increased risk of IA-irAE in a relatively small patient cohort [10]. Although this observation aligns with previous reports of potential OA activation following ICI therapy [11], it is important to note that confounding factors (e.g., body mass index (BMI)) and the small cohort size limited thorough analysis of OA characteristics. In this retrospective study, we systematically evaluate the prevalence and clinical characteristics of preexisting OA among patients who developed *de novo* IA-irAE and compared them with two control groups: ICI-treated patients who developed other irAEs without IA (non-IA irAE) and those who did not develop any irAEs (non-irAE), within a large single-center cohort. We hypothesized that preexisting OA is associated with an increased risk of IA-irAE following ICI therapy.

## PATIENTS AND METHODS

### Patients

This retrospective study was conducted in Mayo Clinic Minnesota to compare the prevalence of OA in ICI-treated cancer patients between January 1, 2015, and June 15, 2025. Patients were adults aged > 18 years with any concomitant malignancy being treated with any ICI including anti-PD-1 blockade, anti-PD-L1 blockade, anti-CTLA-4 blockade as monotherapy or combined therapy. For IA-irAE patients, the clinical diagnosis of IA was made by a consulting rheumatologist and based on inflammatory signs and symptoms of the affected joints. Meanwhile, all the IA developed after the ICI treatment initiation (de novo). The exclusion criteria for IA-irAE patients were: (1) preexisting rheumatic diseases reactivated after ICI treatment, including rheumatoid arthritis, polymyalgia rheumatica, psoriasis arthritis, juvenile rheumatoid arthritis, crystal related arthritis; (2) inflammatory arthritis not related to ICI treatment; (3) ICI treatment duration of fewer than 12 weeks prior to inclusion; (4) other *de novo* rheumatic diseases; (5) only arthralgia without objective inflammatory arthritis; (6) unavailable or incomplete clinical information. Two control groups were included: a non-IA irAE control group in which patients received ICI and developed other types of irAE but not IA, and a non-irAE control group in which patients received ICI but did not develop any types of irAE. After identifying the patients with IA-irAE, frequency matching methods were used to randomly select patients who received ICI of similar age, sex, body mass index (BMI) and smoking status to the IA-irAE case group. These patients were manually reviewed to determine the relevant control group based on the presence or absence of irAE. This study was approved by the Institutional Review Board of Mayo Clinic (IRB 25-006716).

### Data extraction

The electronic medical records were retrospectively reviewed. The data were collected as follows: the demographic information including age, sex, race, BMI, smoking history, family disease history (including OA and AID). Disease-related information including cancer type, cancer stage, ICI type, duration of ICI therapy, irAEs type, malignancy outcomes, time to irAE onset following ICI initiation. Lab test results including immune profile (rheumatoid factor (RF), antinuclear antibody (ANA), anti–cyclic citrullinated peptide antibody (CCP)) were collected in this study. For IA-irAE patients, the following information including IA involved joints, morning stiffness, the erosion on X-ray at any joint sites, the therapeutics, outcomes of IA treatment, the systemic inflammation markers (C-reactive protein (CRP), and erythrocyte sedimentation rate (ESR)), the arthritis severity graded according to the Common Terminology Criteria for Adverse Events (CTCAE) [12], the duration of rheumatology follow-up (defined as the time interval between the first referral to rheumatology and the last rheumatology clinic visit) were also collected. OA history and involved joint sites were recorded for all patients. The diagnosis of OA was based on documented medical history records or on the X-ray of the affected joints that met the Kellgren and Lawrence radiographic criteria [13].

### Statistical analysis

All statistical analyses were performed using GraphPad Prism (version 10.0.0; San Diego, CA, USA) and R Studio software (version 4.5.1). Continuous variables were presented as mean ± standard deviation (SD) or median with 25^th^ -75^th^ percentiles (interquartile range, IQR), whereas categorical variables were summarized as numbers (percentages). For comparisons among three groups, one-way ANOVA was conducted. For comparisons between two groups, Student’s *t* test or Mann–Whitney *U* test was applied for continuous variables. For comparisons of categorical variables, Chi-square test or Fisher’s exact test were used. Logistic regression analyses were conducted to evaluate associations between clinical factors and the development of IA-irAE, with results reported as odds ratios (ORs) and 95% confidence intervals (CIs). A two-sided *p* value < 0.05 was considered statistically significant.

## RESULTS

After initial screening of 14,124 patients, 181 IA-irAE patients were included based on the inclusion and exclusion criteria. In total, this retrospective study included 491 patients receiving ICI treatment, including 181 IA-irAE patients, 140 non-IA irAE patients and 170 non-irAE patients (**Figure 1**).

**Figure 1.**
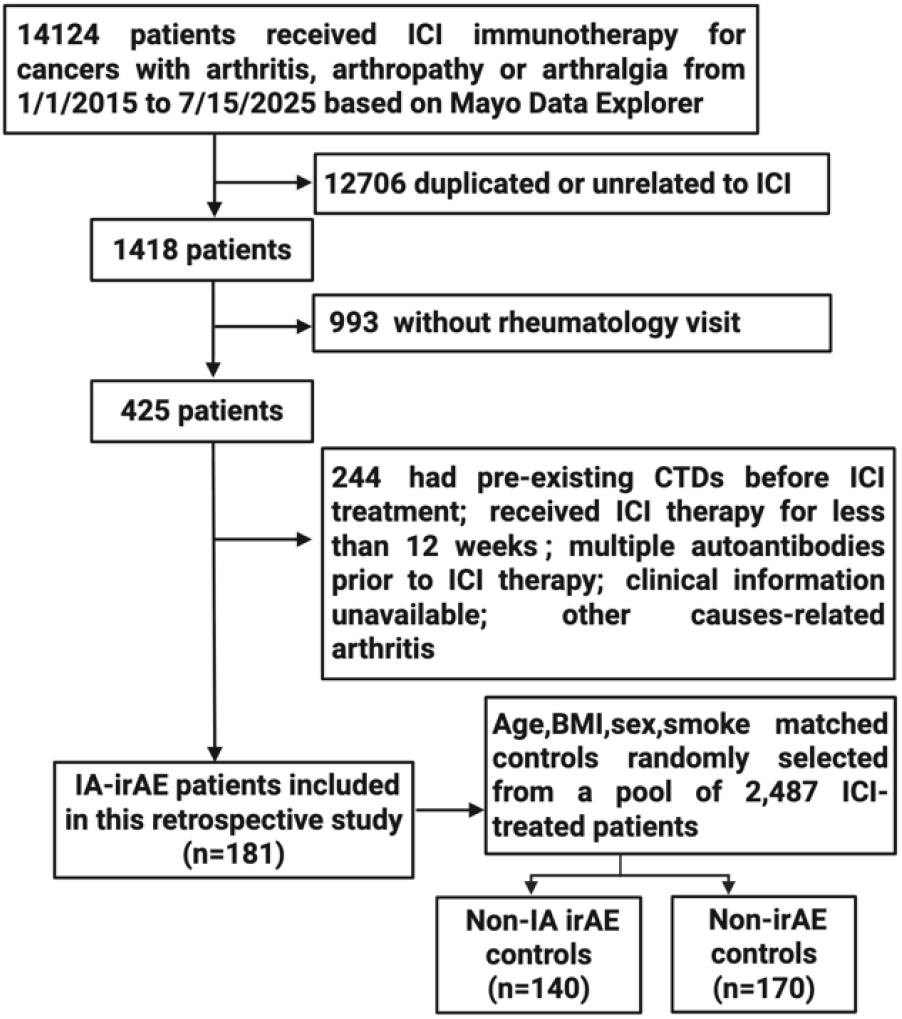
Flow diagram of patients screening

### Comparison of patients’ characteristics among three ICI-treated groups

The mean age of all these patients was 65.6±11.3 and 282 (57%) were male. No significant differences in age, sex distribution, smoking history, BMI, race, cancer stages, AID family history and autoantibody profiles (RF/CCP/ANA) among three groups were observed. Melanoma was the most common cancer in IA-irAE group (55/181, 30%), with a significantly higher prevalence compared with non-IA irAE group (14%, p < 0.001) and non-irAE group (10%, p < 0.001) (**Table 1** and **supplementary Table 1**). Lung cancer was the most common cancer type in both non-IA irAE group (39, 28%) and non-irAE group (56, 33%); however, lung cancer prevalence was not significantly different between 3 groups (p = 0.20) (**Table 1**).

**Table 1.**
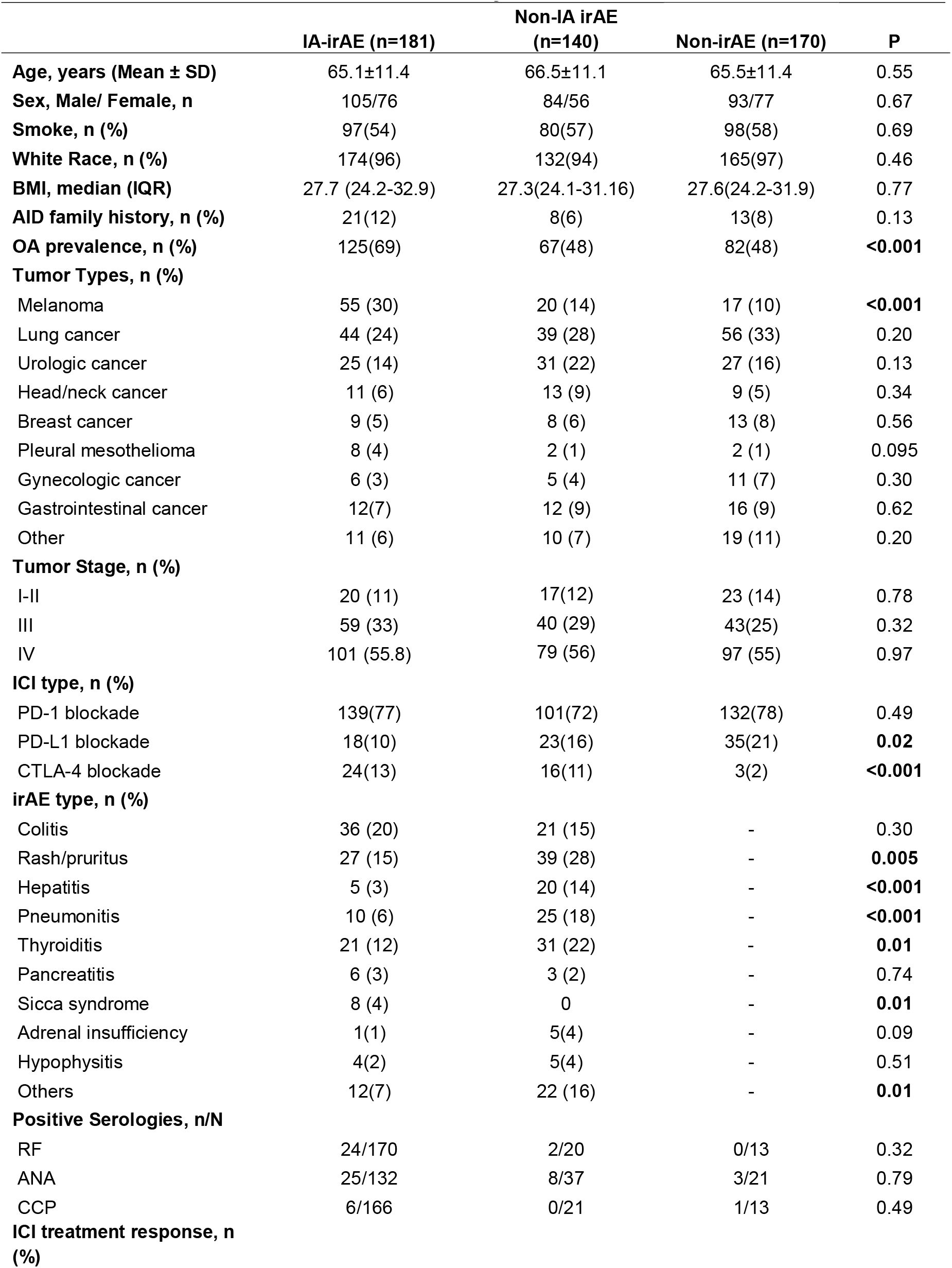
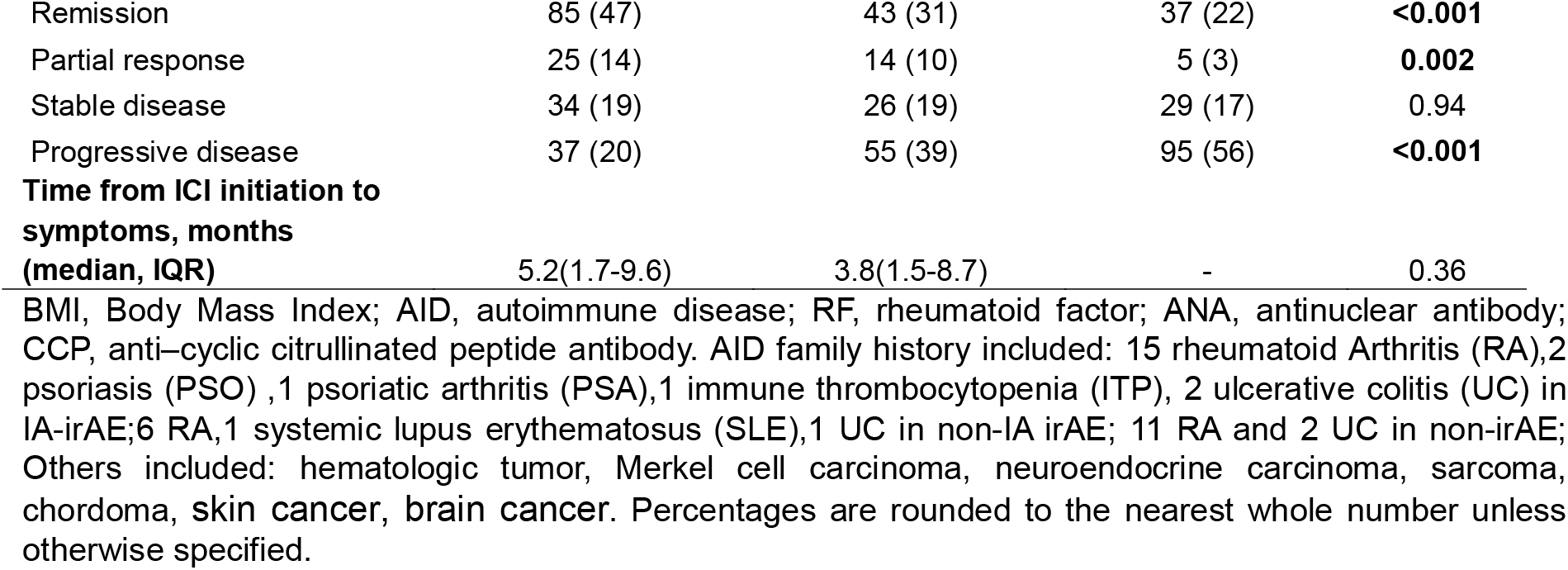
Characteristics of ICI-treated patients included in this retrospective study.

Regarding ICI regimens, a higher proportion of patients in the IA-irAE group received PD-L1 blockade compared with non-irAE group (77% vs 72.1%, p = 0.007, **supplementary Table 1**). In addition, CTLA-4 blockade was more frequently associated with either IA-irAE or non-IA irAE than non-irAE (p < 0.001, **Table 1** and **supplementary Table 1**), consistent with observation that CTLA-4 blockade elicits the highest rate of irAE of any types. PD-1 blockade (including Pembrolizumab, Nivolumab, Cemiplimab and Dostarlimab) were used in 139 (77%) IA-irAE patients, 101 (72%) non-IA irAE patients, and 132 (78%) non-irAE patients; PD-L1 blockade (including Durvalumab, Atezolizumab and Avelumab) in 18(10%) IA-irAE patients, 23(16%) non-IA irAE patients, and 35(21%) non-irAE patients; CTLA-4 blockade (Ipilimumab) in 24 (13%) IA-irAE patients, 16 (11%) non-IA irAE patients, and 3(2%) non-irAE patients. PD-L1 usage was less prevalent in IA-irAE patients (p = 0.02), but more prevalent in non-irAE group (**supplementary Table 1**).

In addition to IA, 96 (53%) patients developed other types of irAE in IA-irAE group. The most common irAE in IA-irAE group was colitis with a rate of 36 (20%) without statistical difference between the two groups. Higher rates of rash/pruritus, hepatitis, pneumonitis and thyroiditis were found in non-IA irAE and non-irAE groups than IA-irAE group (all p < 0.05). Sicca syndrome was more common in IA-irAE compared to non-IA irAE group (p = 0.01). Patients who developed IA-irAE tended to have better cancer outcomes. The remission rate was 85 (47%) in IA-irAE group, higher than 43 (31%) in non-IA irAE group and 37 (22%) in non-irAE group (p < 0.001, **Table 1** and **supplementary Table 1**).

### The prevalence of OA in the three ICI-treated groups

The overall prevalence of OA was 125 (69%) in IA-irAE patients, significantly higher than the 67 (48%) observed in non-IA irAE patients (p < 0.001) and 82 (48%) in non-irAE patients (p < 0.001) (**Figure 2**). A stratified analysis was performed using 65 years of age as the cutoff. Among younger patients (< 65 years), the prevalence of OA was 43 (50%) in IA-irAE group, which was higher than that in non-IA irAE group (15 (25%), p < 0.001) and non-irAE group (29 (38%), p = 0.06). Among older patients (≥ 65 years), the prevalence of OA remained significantly higher in IA-irAE group (79 (83%)) compared with non-IA irAE group (52 (65%), p = 0.008) and non-irAE group (53 (60%), p < 0.001). Together, these findings indicate that OA is more common among ICI-treated patients who developed *de novo* IA-irAE, regardless of age group.

**Figure 2.**
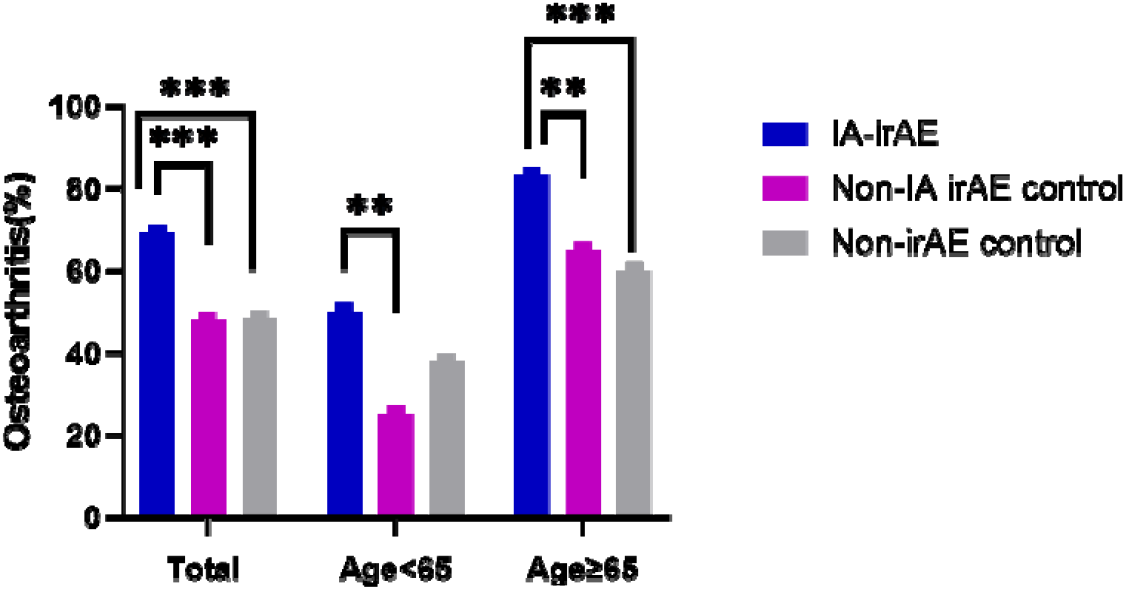
Prevalence of osteoarthritis in the total IA-irAE, non-IA irAE, and non-irAE patients, stratified by age (< 65 years vs. ≥ 65 years). **, p < 0.01; ***, p < 0.001. All p Values were calculated using Fisher’s exact test.

### Subgroup comparison of IA-irAE patients with OA and without OA

The comparison between IA-irAE patients with or without OA was presented in **Table 2**. The mean age of IA-irAE with OA was older than these without OA (p < 0.001) and the BMI of IA-irAE with OA patients demonstrated higher in this subpopulation (p = 0.01). There were no differences in other clinical characteristics including morning stiffness and arthritis severity, or inflammatory markers or immune serologies between two groups (all p > 0.05).

**Table 2.**
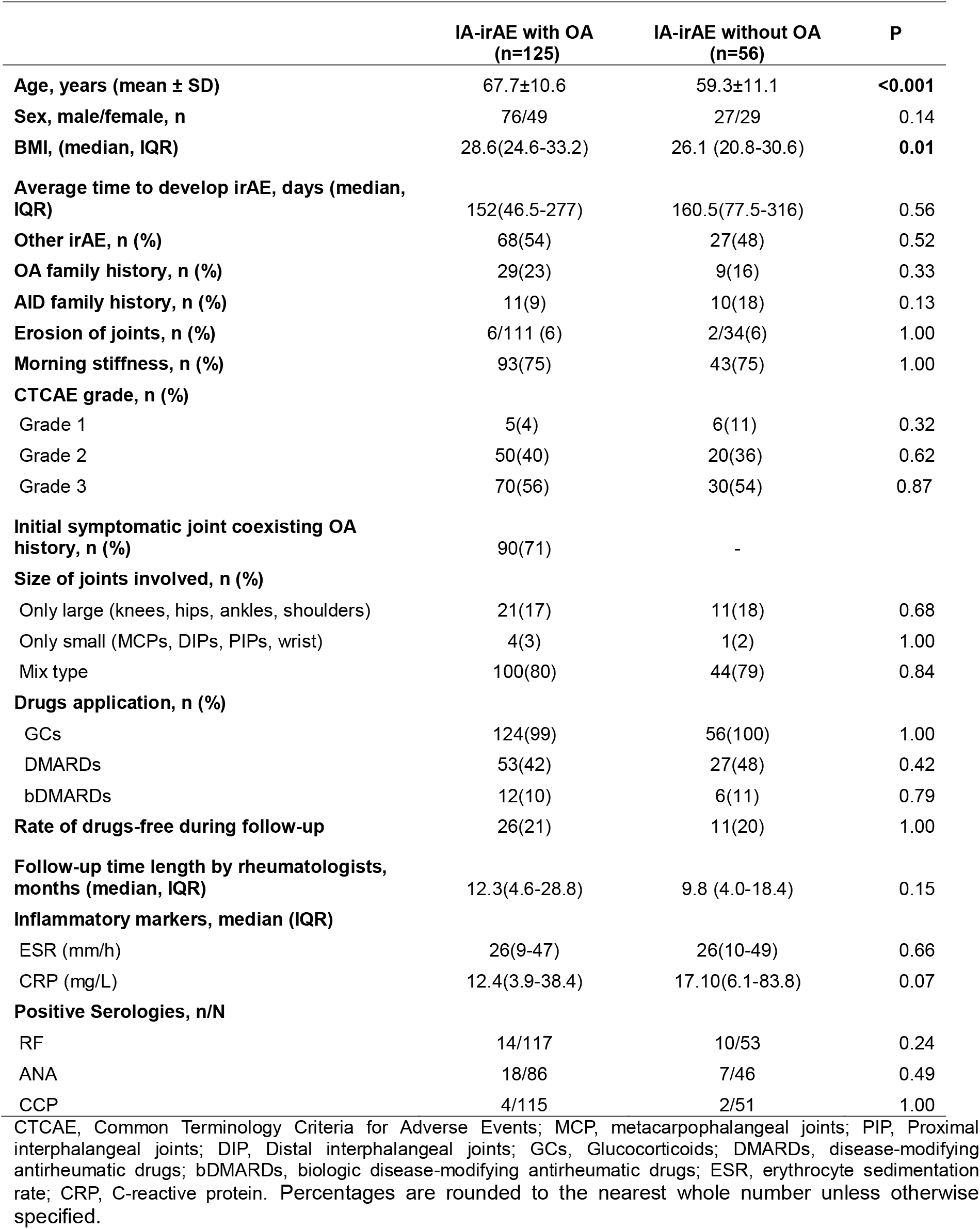
Comparison of IA-irAE patients with or without OA.

|  | IA-irAE with OA<br>(n=125) | IA-irAE without OA<br>(n=56) | P |
| --- | --- | --- | --- |
| **Age, years (mean ± SD)** | 67.7±10.6 | 59.3±11.1 | **<0.001** |
| **Sex, male/female, n** | 76/49 | 27/29 | 0.14 |
| **BMI, (median, IQR)** | 28.6(24.6-33.2) | 26.1 (20.8-30.6) | **0.01** |
| **Average time to develop irAE, days (median, IQR)** | 152(46.5-277) | 160.5(77.5-316) | 0.56 |
| **Other irAE, n (%)** | 68(54) | 27(48) | 0.52 |
| **OA family history, n (%)** | 29(23) | 9(16) | 0.33 |
| **AID family history, n (%)** | 11(9) | 10(18) | 0.13 |
| **Erosion of joints, n (%)** | 6/111 (6) | 2/34(6) | 1.00 |
| **Morning stiffness, n (%)** | 93(75) | 43(75) | 1.00 |
| **CTCAE grade, n (%)** |  |  |  |
| Grade 1 | 5(4) | 6(11) | 0.32 |
| Grade 2 | 50(40) | 20(36) | 0.62 |
| Grade 3 | 70(56) | 30(54) | 0.87 |
| **Initial symptomatic joint coexisting OA history, n (%)** | 90(71) | - |  |
| **Size of joints involved, n (%)** |  |  |  |
| Only large (knees, hips, ankles, shoulders) | 21(17) | 11(18) | 0.68 |
| Only small (MCPs, DIPs, PIPs, wrist) | 4(3) | 1(2) | 1.00 |
| Mix type | 100(80) | 44(79) | 0.84 |
| **Drugs application, n (%)** |  |  |  |
| GCs | 124(99) | 56(100) | 1.00 |
| DMARDs | 53(42) | 27(48) | 0.42 |
| bDMARDs | 12(10) | 6(11) | 0.79 |
| **Rate of drugs-free during follow-up** | 26(21) | 11(20) | 1.00 |
| **Follow-up time length by rheumatologists, months (median, IQR)** | 12.3(4.6-28.8) | 9.8 (4.0-18.4) | 0.15 |
| **Inflammatory markers, median (IQR)** |  |  |  |
| ESR (mm/h) | 26(9-47) | 26(10-49) | 0.66 |
| CRP (mg/L) | 12.4(3.9-38.4) | 17.10(6.1-83.8) | 0.07 |
| **Positive Serologies, n/N** |  |  |  |
| RF | 14/117 | 10/53 | 0.24 |
| ANA | 18/86 | 7/46 | 0.49 |
| CCP | 4/115 | 2/51 | 1.00 |

CTCAE, Common Terminology Criteria for Adverse Events; MCP, metacarpophalangeal joints; PIP, Proximal interphalangeal joints; DIP, Distal interphalangeal joints; GCs, Glucocorticoids; DMARDs, disease-modifying antirheumatic drugs; bDMARDs, biologic disease-modifying antirheumatic drugs; ESR, erythrocyte sedimentation rate; CRP, C-reactive protein. Percentages are rounded to the nearest whole number unless otherwise specified.

### Subgroup comparison of ICI treated patients with OA

Subpopulation analysis was performed among 125 IA-irAE with OA, 67 non-IA irAE with OA and 82 non-irAE with OA patients (**Table 3** and **Supplementary Table 2**). The non-IA irAE with OA patients were older than IA-irAE with OA (p = 0.005) and non-irAE with OA patients (p = 0.006), while no significant differences were observed in comparisons among the other groups. Interestingly, the OA arthritis involvement patterns differed in three groups. In IA-irAE with OA group, patients exhibited more multisite OA, with hand OA being the predominant type (77, 62%), compared to 9 patients (13%) in the IA-irAE with OA group and 11 patients (13%) in the non-irAE with OA group (both p < 0.001). By contrast, knee OA dominated in Non-IA irAE and Non-irAE with OA groups (34 (51%) and 34 (42%), respectively). Other clinical characteristics, inflammatory markers, and immune serologies were indistinguishable between these 3 groups (all p > 0.05).

**Table 3.** Comparison among three ICI-treated cancer patients with OA.

|  | IA-irAE with OA<br>(n=125) | Non-IA irAE with<br>OA (n=67) | Non-irAE with OA<br>(n=82) | P |
| --- | --- | --- | --- | --- |
| **Age, years (mean ± SD)** | 67.7±10.6 | 72.1±9.1 | 67.8± 9.5 | **0.009** |
| **Sex, Male/ Female, n** | 76/49 | 37/30 | 44/38 | 0.55 |
| **BMI, median (IQR)** | 28.6 (24.6-33.2) | 27.7(24.3 -31.1) | 28.0 (24.7-32.1) | 0.63 |
| **Smoke, n (%)** | 74(59) | 42(63) | 50(61) | 0.89 |
| **AID family history, n (%)** | 12(10) | 6(9) | 3(4) | 0.26 |
| **OA family history, n (%)** | 29(23) | 8(12) | 17(21) | 0.10 |
| **OA involvement joints, n (%)** |  |  |  |  |
| Hand | 77(62) | 9 (13) | 11 (13) | **<0.001** |
| Knee | 61(49) | 34 (51) | 34 (42) | 0.46 |
| Hip | 31(25) | 10 (15) | 16 (20) | 0.26 |
| Shoulder | 22(18) | 8 (12) | 14 (17) | 0.57 |
| Feet | 10(8) | 5 (8) | 2 (2) | 0.24 |
| Spine | 16(13) | 5 (8) | 7 (9) | 0.43 |
| Unknown | 1(1) | 9 (13) | 10 (12) | - |
| **Positive Serologies, n/N** |  |  |  |  |
| RF | 6/117 | 1/12 | 0/10 | 0.67 |
| ANA | 18/86 | 2/15 | 3/12 | 0.73 |
| CCP | 4/115 | 0/12 | 1/10 | 0.45 |

Percentages are rounded to the nearest whole number unless otherwise specified.

### Multivariable logistic regression analyses for the risk for IA-irAE

Based on whether patients had IA-irAE, all the 491 ICI-treated patients were classified into 2 groups, 181 IA-irAE patients and 310 non-IA patients. The clinical characteristics were analyzed in **Table 4**. Although CTLA-4 blockade was significantly associated with IA-irAE risk in univariable analysis (OR 2.49, 95%CI 1.32–4.78), its effect could not be independently estimated in the multivariable model due to collinearity with tumor type and other ICI regimens. Other factors including the AID family history (OR 2.03, 95% CI, 1.02–4.05), OA (OR 2.88,95% CI, 1.85–4.52), melanoma (OR 2.63, 95% CI, 1.56–4.47) were all independently associated with IA-irAE development. Surprisingly, PD-L1 blockade was associated with lower odds for IA-irAE development (OR 0.30,95% CI, 0.12–0.74) in univariable analyses with the caveat that the number of patients treated with PD-L1 inhibitor was much smaller compared to PD-1 inhibitor (**Table 1**).

**Table 4.**
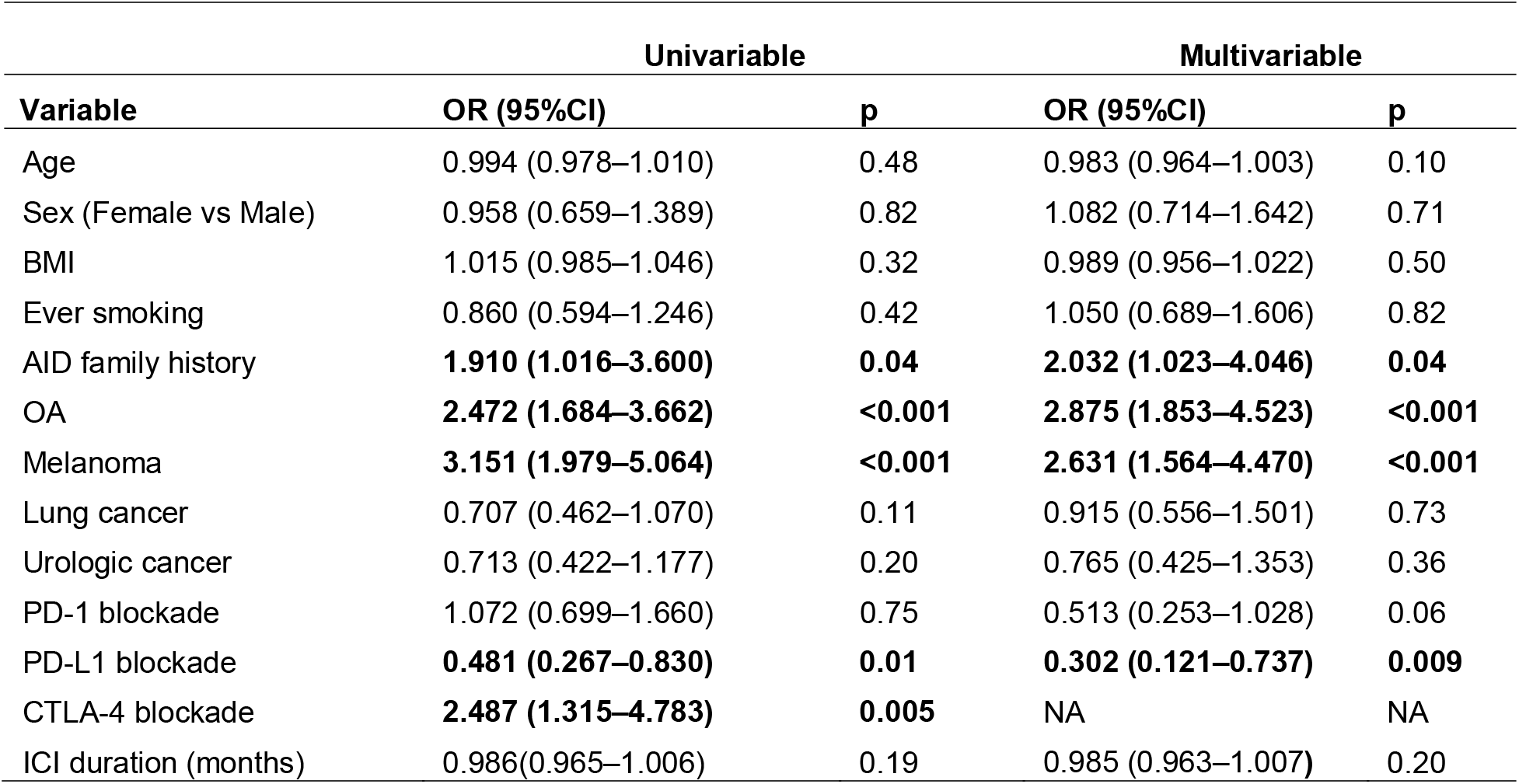
Univariable and multivariable logistic regression analyses for risk factors of IA-irAE.

## DISCUSSION

This work represents the largest study focused on the prevalence of OA in *de novo* IA-irAE patients. We conclude that OA is a real risk factor for IA-iAE. The overall prevalence of OA was 69% in IA-irAE patients, 48% in non-IA irAE patients and 48% in non-irAE patients. The prevalence remained consistently higher in the IA-irAE group compared with the other two groups, regardless of age. We further identify that melanoma, family history of AID, and CTLA-4 blockade are risk factors for IA-irAE development. Among ICI-treated patients with OA, the pattern of joint involvement differed across subgroups, with hand OA being more predominant in those who developed IA-irAEs.

Radiographic OA has been reported in 39.1%–66.6% of patients with IA-irAE in two prior studies, including a multicenter observational study and a single-center experience [14, 15]. However, none of these studies included control groups. More recently, Matthieu and colleagues reported that 84.9% of IA-irAE patients had a history of OA and 89.3% had radiographic OA, and the rates were substantially higher than those observed in other ICI-treated populations [10]. However, the study might be confounded by differences of BMI, a well-established risk factor for OA, because their IA-irAE group had higher BMI than the control groups. Furthermore, all these studies were limited by small sample sizes, with fewer than 40 IA-irAE patients included. Our study examined the largest cohort of *de novo* IA-irAE patients with OA to date, which specifically focused on the prevalence and characteristics of OA in *de novo* IA-irAE with control groups matched for BMI, age and sex. Consistent with the previous study, OA was more common in IA-irAE patients than in non-IA irAE or non-irAE patients. Furthermore, among patients with OA, we observed distinct OA involvement patterns across the three ICI-treated groups. IA-irAE patients were characterized by a predominance of hand OA and a polyarticular distribution, distinguishing them from the control groups. Seronegative RA-like polyarthritis has been reported as the most common type of IA-irAE [4, 16, 17]. In line with this observation, our two previous studies demonstrated that 65%–75% of IA-irAE patients presented with polyarticular arthritis, most commonly involving both small and large joints [18, 19]. The high prevalence of hand OA observed in IA-irAE therefore is unlikely to be a coincidental finding. In a multicenter retrospective study of 26 patients who developed symptomatic OA without objective joint inflammation after ICI initiation (ICI-aOA), small-joint involvement was observed in only 25% of cases [11], a distribution clearly distinct from that observed in IA-irAE [16]. Since low-grade inflammatory OA and erosive OA have been proposed [20], OA is no longer considered a purely non-inflammatory condition, but rather a heterogeneous disease spectrum with distinct inflammatory phenotypes. Together, these findings suggest that pre-existing hand OA may be predisposed to the development of polyarticular IA-irAE, rather than reflecting a nonspecific consequence of ICI exposure.

To date, relatively few clinical characteristics have been consistently associated with the development of rheumatic irAEs. Melanoma and genitourinary cancer were found to be the risk factors of rheumatic irAEs and *de novo* IA-irAE in a large case– control study [8]. However, other potentially relevant clinical variables, such as BMI, smoking status, and family history of AID, were not included in the analysis. Machine learning analyses showed that patients with melanoma and renal cell carcinoma were more likely to develop IA-irAE compared with patients with lung cancer [21]. Melanoma is associated with an approximately 2.6-fold increased risk of IA-irAE, while urologic cancer is not identified in our study. Sex, Age and BMI are well-recognized risk factors for OA [13]. In addition, elevated BMI has been associated with an increased risk of irAEs [22]. Our study minimizes the potential impact of these confounding factors on OA prevalence. Although preexisting AIDs have been associated with an increased risk of irAEs [23, 24], we exclude patients with preexisting AIDs from the present study and instead focus on *de novo* IA-irAE. Evidence supporting the role of family history of AID remains scarce and is largely derived from expert opinion rather than primary data. We find a family history of AID is associated with the development of IA-irAE when compared with ICI-treated controls without IA, thus providing novel real-world evidence supporting a role for autoimmune predisposition in the development of IA-irAE. The risk profiles of different ICI classes for irAEs have been well summarized in several meta-analyses [3, 25]. Notably, our data is consistent with previous studies that CTLA-4 inhibition is associated with higher incidence of irAE [25]. While our data suggests that PD-L1 blockade is associated with reduced odds of IA-irAE develop, the sample size of IA-irAE patients treated with PD-L1 inhibitor is relatively small and different PD-L1 inhibitors may be associated with different rate of irAE development[8]. Thus, the association between PD-L1 blockade and IA-irAE development awaits further investigation.

Several limitations of this study should be acknowledged. First, this is a retrospective study focused primarily on clinical risk factors of interest, particularly OA, and therefore could not account for other potentially relevant variables, including detailed laboratory evaluations such as lymphocyte subsets or serological biomarkers. Second, this study includes only patients evaluated by rheumatologists. As a result, cases of IA-irAE managed without specialty referral could be underrecognized, potentially leading to an underestimation of the true prevalence of IA-irAE with or without OA. Accordingly, future prospective studies incorporating immunologic profiling and broader case ascertainment are warranted.

In conclusion, as the largest study to evaluate the prevalence of OA in IA-irAE patients, we identify hand OA as a joint-level susceptibility shaping the clinical phenotype of IA-irAE. Additionally, melanoma, a family history of AID were other risk factors for IA-irAE, while the use of anti PD-L1 antibody is associated with lower observed odds of IA-irAE.

## Data Availability

All data produced in the present study are available upon reasonable request to the authors.

## ACKNOWLEDGMENT

We thank Dr. Selene Rubino for her help with Mayo Data Explorer.

## AUTHOR CONTRIBUTIONS

All authors approved the final version for publication. Dr. Zeng retained full access to all the data in the study and takes responsibility for the integrity of the data and the accuracy of the data analysis.

### Study conception and design

Chen, Thanarajasingam, Crowson, Zeng

### Acquisition of data

Chen, Zhu, Crowson.

### Analysis and interpretation of data

Chen, Crowson, Zhu, Zhang, Zeng

### Manuscript draft and editing

Chen, Zhu, Crowson, Zeng

