## Supplementary Table 1 and 2 for "The association of osteoarthritis with the risk of de novo inflammatory arthritis in patients receiving immune checkpoint inhibitors: a retrospective study"

| **Supplementary Table 1. Characteristics of ICI-treated patients included in this retrospective study** | | | | | | | | | | | | |
| --- | --- | --- | --- | --- | --- | --- | --- | --- | --- | --- | --- | --- |
|  | | **IA-irAE (n=181)** | | **Non-IA irAE (n=140)** | | **Non-irAE (n=170)** | | **P** | | **P1** | | **P2** |
| **Age, years (Mean ± SD)** | | 65.1±11.4 | | 66.5±11.1 | | 65.5±11.4 | | 0.55 | | 0.28 | | 0.76 |
| **Sex, Male/ Female, n** | | 105/76 | | 84/56 | | 93/77 | | 0.67 | | 0.76 | | 0.59 |
| **Smoke, n (%)** | | 97(54) | | 80(57) | | 98(58) | | 0.69 | | 0.56 | | 0.45 |
| **White Race, n (%)** | | 174(96) | | 132(94) | | 165(97) | | 0.46 | | 0.44 | | 0.77 |
| **BMI, median (IQR)** | | 27.7 (24.2-32.9) | | 27.3(24.1-31.2) | | 27.6(24.2-31.9) | | 0.77 | | 0.49 | | 0.71 |
| **AID family history, n (%)** | | 21(12) | | 8(6) | | 13(8) | | 0.13 | | 0.08 | | 0.15 |
| **OA prevalence, n (%)** | | 125(69) | | 67(48) | | 82(48) | | **<0.001** | | **<0.001** | | **<0.001** |
| **Tumor Types, n (%)** | |  | |  | |  | |  | |  | |  |
| Melanoma | | 55 (30) | | 20 (14) | | 17 (10) | | **<0.001** | | **<0.001** | | **<0.001** |
| Lung cancer | | 44 (24) | | 39 (28) | | 56 (33) | | 0.20 | | 0.52 | | 0.08 |
| Urologic cancer | | 25 (14) | | 31 (22) | | 27 (16) | | 0.13 | | 0.55 | | 0.65 |
| Head/neck cancer | | 11 (6) | | 13 (9) | | 9 (5) | | 0.34 | | 0.29 | | 0.82 |
| Breast cancer | | 9 (5) | | 8 (6) | | 13 (8) | | 0.56 | | 0.81 | | 0.38 |
| Pleural mesothelioma | | 8 (4) | | 2 (1) | | 2 (1) | | 0.10 | | 0.20 | | 0.11 |
| Gynecologic cancer | | 6 (3) | | 5 (4) | | 11 (7) | | 0.30 | | 1.00 | | 0.22 |
| Gastrointestinal cancer | | 12(7) | | 12 (9) | | 16 (9) | | 0.62 | | 0.53 | | 0.43 |
| Other | | 11 (6) | | 10 (7) | | 19 (11) | | 0.19 | | 0.82 | | 0.13 |
| **Tumor Stage, n (%)** | |  | |  | |  | |  | |  | |  |
| I-II | | 20 (11) | | 17(12) | | 23 (14) | | 0.78 | | 0.86 | | 0.52 |
| III | | 59 (33) | | 40 (29) | | 43(25) | | 0.32 | | 0.47 | | 0.16 |
| IV | | 101 (56) | | 79 (56) | | 97 (55) | | 0.97 | | 1.00 | | 0.83 |
| Others | | 1(1) | | 4(3) | | 7(4) | | 0.07 | | 0.17 | | **0.03** |
| **ICI type, n (%)** | |  | |  | |  | |  | |  | |  |
| PD-1 blockade | | 139(77) | | 101(72) | | 132(78) | | 0.49 | | 0.37 | | 0.90 |
| PD-L1 blockade | | 18(10) | | 23(16) | | 35(21) | | **0.02** | | 0.09 | | **0.007** |
| CTLA-4 blockade | | 24(13) | | 16(11) | | 3(2) | | **<0.001** | | 0.73 | | **<0.001** |
| **irAE type, n (%)** | |  | |  | |  | |  | |  | |  |
| Colitis | | 36 (20) | | 21 (15) | | - | | - | | 0.30 | | - |
| Rash/pruritus | | 27 (15) | | 39 (28) | | - | | - | | **0.005** | | - |
| Hepatitis | | 5 (3) | | 20 (14) | | - | | - | | **<0.001** | | - |
| Pneumonitis | | 10 (6) | | 25 (18) | | - | | - | | **<0.001** | | - |
| Thyroiditis | | 21 (11) | | 31 (22) | | - | | - | | **0.01** | | - |
| Pancreatitis | | 6 (3) | | 3 (2) | | - | | - | | 0.74 | | - |
| Sicca syndrome | | 8 (4) | | 0 | | - | | - | | **0.01** | | - |
| Adrenal insufficiency | | 1(1) | | 5(4) | | - | | - | | 0.09 | | - |
| Hypophysitis | | 4(2.2) | | 5(4) | | - | | - | | 0.51 | | - |
| Others | | 12(7) | | 22 (1) | | - | | - | | **0.01** | | - |
| **Positive Serologies, n/N** | |  | |  | |  | |  | |  | |  |
| RF | | 24/170 | | 2/20 | | 0/13 | | 0.32 | | 1.00 | | 0.22 |
| ANA | | 25/132 | | 8/37 | | 3/21 | | 0.79 | | 0.82 | | 0.77 |
| CCP | | 6/166 | | 0/21 | | 1/13 | | 0.49 | | 1.00 | | 0.42 |
| **ICI treatment response, n (%)** | |  | |  | |  | |  | |  | |  |
| Remission | | 85 (47) | | 43 (31) | | 37 (22) | | **<0.001** | | **0.004** | | **<0.001** |
| Partial response | | 25 (14) | | 14 (10) | | 5 (3) | | **0.002** | | **<0.001** | | 0.39 |
| Stable disease | | 34 (19) | | 26 (19) | | 29 (17) | | 0.94 | | 0.89 | | 0.89 |
| Progressive disease | | 37 (20) | | 55 (39) | | 95 (56) | | **<0.001** | | **<0.001** | | 0.79 |
| **Time from ICI initiation to symptoms, months (median, IQR)** | | 5.2(1.7-9.6) | | 3.8(1.5-8.7) | | - | | - | | 0.36 | | - |
| BMI, Body Mass Index; AID, autoimmune disease; RF, rheumatoid factor; ANA, antinuclear antibody; CCP, anti–cyclic citrullinated peptide antibody. AID family history included: 15 rheumatoid Arthritis (RA),2 psoriasis (PSO) ,1 psoriatic arthritis (PSA),1 immune thrombocytopenia (ITP), 2 ulcerative colitis (UC) in IA-irAE;6 RA,1 systemic lupus erythematosus (SLE),1 UC in non-IA irAE; 11 RA and 2 UC in non-irAE; Others included: hematologic tumor, Merkel cell carcinoma, neuroendocrine carcinoma, sarcoma, chordoma, skin cancer, brain cancer; P, comparison among the three groups; P1, comparison between the IA-irAE and non-IA irAE groups; P2, comparison between the IA-irAE and non-irAE groups. Percentages are rounded to the nearest whole number unless otherwise specified.  **Supplementary Table 2. Comparison among three ICI-treated cancer patients with OA** | | | | | | | | | | | | |
|  | **IA-irAE with OA (n=125)** | | **Non-IA irAE with OA (n=67)** | | **Non-irAE with OA (n=82)** | | **P** | | **P1** | | **P2** | |
| **Age, years (mean ± SD)** | 67.7±10.6 | | 72.1±9.1 | | 67.8± 9.5 | | **0.009** | | **0.008** | | 0.86 | |
| **Sex, Male/ Female, n** | 76/49 | | 37/30 | | 44/38 | | 0.55 | | 0.54 | | 0.32 | |
| **BMI, median (IQR)** | 28.6 (24.6-33.2) | | 27.7(24.3 -31.0) | | 28.0 (24.7-32.1) | | 0.63 | | 0.35 | | 0.68 | |
| **Smoke, n (%)** | 74(59) | | 42(63) | | 50(61) | | 0.89 | | 0.76 | | 0.89 | |
| **AID family history, n (%)** | 12(10) | | 6(9) | | 3(4) | | 0.26 | | 1.00 | | 0.17 | |
| **OA family history, n (%)** | 29(23) | | 8(12) | | 17(21) | | 0.10 | | **0.04** | | 0.50 | |
| **OA involvement joints, n (%)** | | |  | |  | |  | |  | |  | |
| Hand | 77(62) | | 9 (13) | | 11 (13) | | **<0.001** | | **<0.001** | | **<0.001** | |
| Knee | 61(49) | | 34 (51) | | 34 (42) | | 0.46 | | 0.88 | | 0.32 | |
| Hip | 31(25) | | 10 (15) | | 16 (20) | | 0.26 | | 0.14 | | 0.40 | |
| Shoulder | 22(18) | | 8 (12) | | 14 (17) | | 0.57 | | 0.41 | | 1.00 | |
| Feet | 10(8) | | 5 (8) | | 2 (2) | | 0.24 | | 1.00 | | 0.13 | |
| Spine | 16(13) | | 5 (8) | | 7 (9) | | 0.43 | | 0.34 | | 0.50 | |
| Unknown | 1(1) | | 9 (13) | | 10 (12) | | - | | - | | - | |
| **Positive Serologies, n/N** | | |  | |  | |  | |  | |  | |
| RF | 6/117 | | 1/12 | | 0/10 | | 0.67 | | 0.50 | | 1.00 | |
| ANA | 18/86 | | 2/15 | | 3/12 | | 0.73 | | 0.73 | | 0.72 | |
| CCP | 4/115 | | 0/12 | | 1/10 | | 0.45 | | 1.00 | | 0.35 | |

P, comparison among the three groups; P1, comparison between the IA-irAE and non-IA irAE groups; P2, comparison between the IA-irAE and non-irAE groups. Percentages are rounded to the nearest whole number unless otherwise specified.
